# *Post-Mortem* validation of *in vivo* 18kDa Translocator Protein (TSPO) PET as a microglial biomarker

**DOI:** 10.1101/2024.07.15.24309178

**Authors:** Sasvi S. Wijesinghe, James B. Rowe, Hannah D. Mason, Kieren S. J. Allinson, Reuben Thomas, Davi Vontobel, Tim D. Fryer, Young T. Hong, Mehtap Bacioglu, Maria Grazia Spillantini, Jelle Van den Ameele, John T. O’Brien, Sanne Kaalund, Maura Malpetti, Annelies Quaegebeur

## Abstract

Neuroinflammation is a feature of many neurodegenerative diseases, and can be quantified *in vivo* by PET imaging with radioligands for the translocator protein (TSPO, e.g. [^11^C]-PK11195). TSPO radioligand binding correlates with clinical severity and predicts clinical progression. However, the cellular substrate of altered TSPO binding is controversial and requires neuropathological validation.

We used progressive supranuclear palsy (PSP) as a demonstrator condition, to test the hypothesis that [^11^C]-PK11195 PET reflects microglial changes. We included people with PSP-Richardson’s syndrome who had undergone [^11^C]-PK11195 PET in life. In *post-mortem* brain tissue from the same participants we characterised cell-type specific TSPO expression with double-immunofluorescence labelling and quantified microgliosis in eight cortical and eleven subcortical regions with CD68 immunohistochemistry.

Double-immunofluorescence labelling for TSPO and cell markers showed TSPO expression in microglia, astrocytes, and endothelial cells. Microglial TSPO expression was higher in donors with PSP compared to controls, which was not the case for astrocytic TSPO expression. There was a significant positive correlation between regional [^11^C]-PK11195 binding potential *ante-mortem* and the density of *post-mortem* CD68+ phagocytic microglia, as well as microglial TSPO expression.

We conclude that [^11^C]-PK11195 binding *in vivo* is driven by microglia and can be interpreted as a biomarker of microglia-mediated neuroinflammation in tauopathies.

## Introduction

Neuroinflammation has emerged as an important pathological feature of multiple neurodegenerative diseases, alongside accumulation of misfolded protein aggregates, synapse loss and neuronal death^1^. Microglia have gained particular interest as a dominant part of the central neuroinflammatory response, associated with disease severity and progression^2,3^. Given the potential for prognostication and therapeutic targeting of inflammation, biomarkers that can quantify and localise the inflammatory response *in vivo* are of high importance^4^.

Several measures of neuroinflammation have emerged using PET imaging, with radioligands that bind to the 18kDa translocator protein (TSPO), such as [11C]-PK11195^5^. TSPO PET has been used to visualise neuroinflammatory changes in the brain in patients living with neurodegenerative diseases. TSPO binding has been reported to increase in Alzheimer’s disease (AD), Parkinson’s disease, frontotemporal dementias, dementia with Lewy bodies, and progressive supranuclear palsy (PSP)^6–15^. The regional distribution of increased TSPO radioligand binding is disease specific, for example, being highest in temporoparietal regions in AD versus pallidum, midbrain, and frontal cortex in PSP ^6,16,17^. The disease-specific regional binding relates to clinical severity and predicts clinical progression^3,6^. Whilst baseline imaging is prognostic, longitudinal increases in the TSPO are also observed^18,19^.

There remain controversies in the interpretation of TSPO PET. The rationale of TSPO radioligands as biomarkers for neuroinflammation is based largely on autoradiography and animal and cell culture studies. Early studies correlated TSPO autoradiography to microglial staining in neurotoxin treated tissue and preclinical mouse models of AD^20,21^. Antibody-based primate and mouse studies corroborated these findings, and confirmed the correlation of CD68+ microglial staining to TSPO markers when treated with neuroinflammatory agents such as lipopolysaccharides (LPS)^22,23^. However, it has been challenged whether TSPO reflects microglia specifically, and their reactivity or density in brain tissue. For example, TSPO expression does not always increase in response to inflammatory or neurotoxic stimuli^24,25^, and in AD and multiple sclerosis *post-mortem* tissue studies, increased TSPO expression has been observed in endothelium and astrocytes^26,27^. This prompts the question as to which cell-types contribute to the disease-driven changes observed by TSPO PET imaging.

This study had two principal aims: (i) to test the hypothesis that TSPO elevation in tauopathies is microglial-specific, rather than driven by astrocytes, and (ii) to test the hypothesis that *ante-mortem* TSPO PET imaging correlates with regional and individual *post-mortem* differences in microglia. We used PSP as a demonstrator tauopathy, because of its high clinicopathological correlation, prognostic relevance of increased TSPO radioligand binding, and short disease course with a short timeframe between PET and death. Confirmation of these hypotheses would support the interpretation of TSPO PET (specifically [^11^C]-PK11195) as a microglial neuroinflammatory biomarker in primary tauopathies.

## Materials and methods

Eight people with PSP (with *ante-mortem* diagnosis of PSP-Richardson’s syndrome) who underwent [^11^C]-PK11195 PET during life^15,17^ donated their brain to the Cambridge Brain Bank. Neuropathological diagnosis of PSP was confirmed in all cases. Their demographics are summarised in Table 1. A control group consisted of 3 age-matched neurologically healthy controls with minimal to mild age-related pathology.

**Table 1:** Demographics, clinical and pathological data of PSP donors.

| **Case No** | **Gender** | **Disease Duration (years)** | **Pathology (Stage)** | **PET-to-death time (months)** | **PSP Rating Scale at PET** |
| --- | --- | --- | --- | --- | --- |
| 1 | Female | 6.2 | PSP (5) | 30 | 46 |
| 2 | Male | 5.4 | PSP (5) | 28 | 33 |
| 3 | Female | 8.8 | PSP (2) | 49 | 46 |
| 4 | Female | 16.3 | PSP (4) | 6 | 74 |
| 5 | Male | 6.3 | PSP (4) | 23 | 54 |
| 6 | Female | 5.5 | PSP (3) | 34 | 29 |
| 7 | Male | 6.0 | PSP (3) | 18 | 39 |
| 8 | Female | 5.3 | PSP (5) | 24 | 38 |

*PSP: progressive supranuclear palsy*

We used double-immunofluorescence labelling in formalin-fixed paraffin-embedded tissue sections to visualise cell-type specific expression of TSPO in astrocytes (GFAP), microglia (IBA1) and endothelium (CD31) in posterior frontal lobe tissue (Brodmann area 6), followed by Leica© SPE Confocal Microscopy for high magnification images and z-stacks. DAB-based immunohistochemical staining identified expression of CD68 of eight cortical and eleven subcortical areas. Whole-slide images were acquired by an Aperio AT2 whole slide scanner (Leica) at x40 magnification for immunohistochemistry slides, and a Zeiss Axioscan Z1 Slidescanner at x40 for immunofluorescence slides. QuPath quantified CD68+ staining through pixel-classification based analysis in grey and white matter differentiated Regions of Interest (ROIs). Area fraction and co-localisation analysis of the IBA1/TSPO and GFAP/TSPO whole slide-scans was performed using a colour-thresholding pipeline in ImageJ.

Mann-Whitney tests compared control and PSP datasets. The Kruskal-Wallis rank sum test and Dunn’s post-hoc test were applied to the grey matter/white matter comparisons for IBA1-TSPO and GFAP-TSPO area fraction analysis. A linear mixed effects model tested the association between *in vivo* [^11^C]-PK11195 binding potential (BP_ND_) and CD68+ microglia quantification across all regions. Spearman’s correlation analyses were performed to assess the association between the *in vivo* [^11^C]-PK11195 BP_ND_ and TSPO-IBA1 co-localisation and TSPO-GFAP co-localisation area in frontal lobe.

*Please refer to Supplementary Material for detailed Materials and Methods*.

## Results

### TSPO showed a multicellular expression profile in *post-mortem* brain tissue

To investigate the cellular substrate of [^11^C]-PK11195 binding, we characterised the TSPO expression profile in human *post-mortem* tissue. Immunohistochemical staining revealed ubiquitous expression of TSPO across white and grey matter of the frontal lobe with a strong expression in the vasculature (Figure 1A). In addition, non-vascular cellular expression was observed, with staining morphology suggestive of a glial origin.

**Figure 1:**
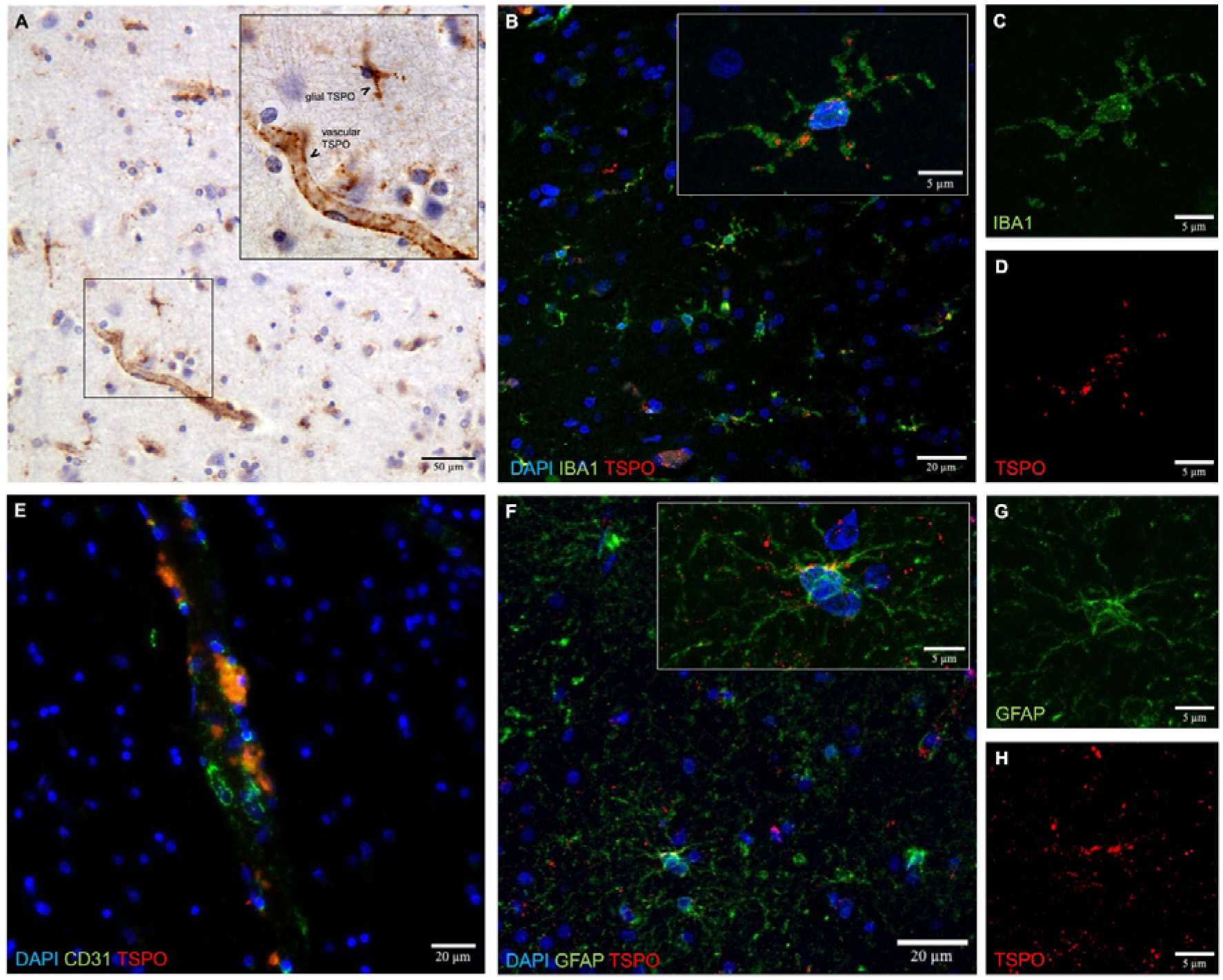
Multicellular expression profile of TSPO in human *post-mortem* brain. (**A**) Immunohistochemistry images of TSPO in frontal lobe tissue. **(B)** Representative immunofluorescence images of TSPO (red), the microglial marker IBA1 (green), and DAPI (blue). **(C, D)** Insets from (B) showing individual channels of **(C)** microglia (IBA1) and **(D)** TSPO staining. **(E)** Representative immunofluorescence images of TSPO and the endothelial marker CD31. **(F)** Representative immunofluorescence images of TSPO (red), the astrocytic marker GFAP (green), and DAPI (blue). **(G, H)** Insets from (F) showing individual channels of **(G)** astrocytes (GFAP) and **(H)** TSPO staining.

To determine the cell-type specific expression of TSPO in more detail, immunofluorescence labelling for TSPO together with cell-type markers (IBA1 for microglia, GFAP for astrocytes and CD31 for endothelial cells) was carried out. IBA1-TSPO co-staining revealed TSPO expression in a large proportion of the IBA1+ microglia, showing a punctate TSPO pattern in microglial soma and processes across white and grey matter (Figure 1B-D). CD31-TSPO co-staining showed abundant expression of TSPO in nearly all endothelial cells, and this was observed across capillaries, arterioles, venules, arteries and veins, as expected from the immunohistochemical observations (Figure 1E). GFAP-TSPO co-staining demonstrated sparse expression of TSPO in astrocytes, with only rare GFAP+ astrocytes showing co-localisation with TSPO staining (Figure 1F-H).

### Quantification of cell-type specific expression revealed a microglia-specific increase of TSPO in PSP

Next, we investigated which cell type drives the increase in TSPO radioligand binding in PSP. Since PSP lacks a significant vascular contribution and the cellular substrate of the TSPO signal is mainly contested between the glial cell types^25^, we focused our analysis on astrocytic and microglial expression. We quantified the area of IBA1-TSPO co-localisation (reflective of microglial TSPO expression) and area of GFAP-TSPO co-localisation (reflective of astrocytic TSPO expression) in posterior frontal lobe (BA6) tissue from neuropathologically confirmed PSP donors who had undergone [^11^C]-PK11195 PET during life^15,17^, and control donors.

There was a significant increase in the microglial TSPO expression in PSP versus controls (IBA1-TSPO co-localisation area fraction in controls: 0.046 ± 0.030%, PSP: 0.460 ± 0.246%, *p* < 0.05; Figure 2A). Comparison of microglial TSPO expression in white (WM) and grey matter (GM) revealed an increase in both compartments in PSP versus controls, although this was more pronounced and statistically significant in white matter (IBA1-TSPO co-localisation area fraction in control WM: 0.089 ± 0.062%, PSP WM: 0.850 ± 0.441%, *p* < 0.01, control GM: 0.042 ± 0.033%, PSP GM: 0.256 ± 0.172%, *p* = ns; Figure 2B).

**Figure 2:**
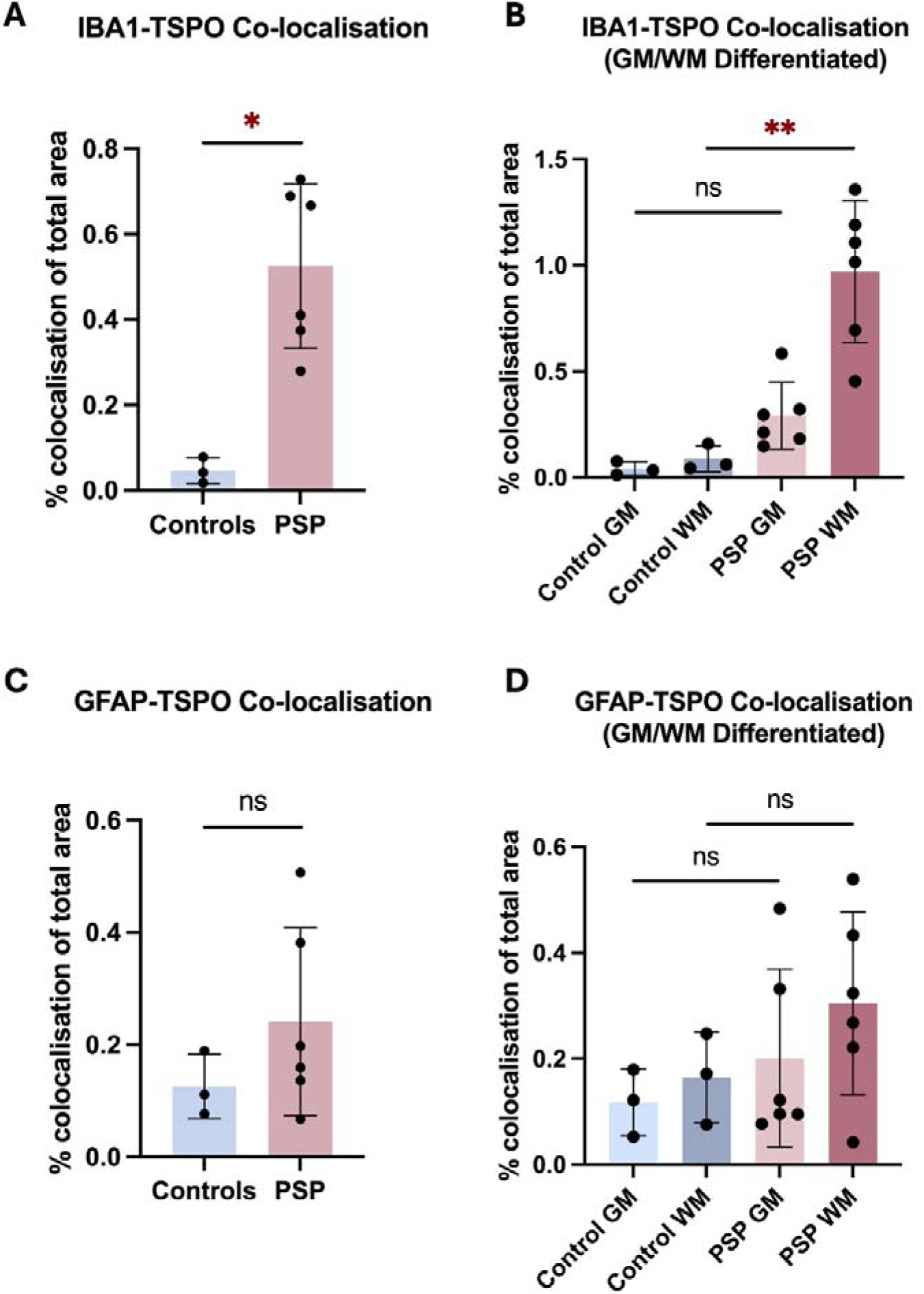
The cell-type specific localisation of TSPO in PSP vs controls. (**A, C**) Area fraction quantification of TSPO and (A) IBA1 (microglia) or (C) GFAP (astrocyte) co-localisation (Mann-Whitney Test; *p* < 0.05; ns = not significant; graph depicts mean ± SD). **(B, D)** Area fraction quantification of TSPO and (B) IBA1 (microglia) or (D) GFAP (astrocyte) co-localisation in the grey (GM) and white matter (WM) compartments (Kruskal-Wallis Rank Sum Test; PSP GM vs WM, *p* < 0.01; ns = not significant; graph depicts mean ± SD).

Astrocytic TSPO expression did not differ significantly between patients versus controls, in frontal lobe as a whole (GFAP-TSPO co-localisation area fraction in controls: 0.126 ± 0.057%, PSP: 0.217 ± 0.166%, *p* = ns; Figure 2C) or in white and grey matter compartments (GFAP-TSPO co-localisation area fraction in control WM: 0.165 ± 0.086%, PSP WM: 0.271 ± 0.181%, *p* = ns, control GM: 0.118 ± 0.063%, PSP GM: 0.182 ± 0.161%, *p* = ns; Figure 2D).

This data suggests that the disease-related TSPO binding in PSP is predominantly driven by microglia, rather than astrocytes, in particular microglia in white matter.

### *Post-mortem* microgliosis and microglial TSPO expression correlated with *ante-mortem* TSPO radioligand binding

To confirm that TSPO radioligand binding determined with PET in PSP reflects microglial reactivity, we assessed the regional association between histologically-determined microgliosis in *post-mortem* brain tissue and [^11^C]-PK11195 binding potential (BP_ND_) from the same PSP donors during life^15,17^. “Phagocytic” microglia were quantified using CD68 immunohistochemistry across 8 cortical and 11 subcortical regions. Highest area fractions of CD68+ microglia were seen across the cortical white matter regions and subcortical regions (Figure 3A). [^11^C]-PK11195 BP_ND_ was calculated for each individual donor for each region (Figure 3B). The model comparison of three linear mixed effects models identified one with a fixed term for the effect of [^11^C]-PK11195 BP_ND_ on CD68 area fraction, and random intercept for individual patients (Δ*chi-square (1)* = 4.47, *p* = 0.0345), but not random slope (Δ*chi-square (1)* = 1.89, *p* = 0.39). The optimal model confirmed the significant positive association between *in vivo* TSPO radioligand binding and the area fraction of *post-mortem* CD68-positive ‘phagocytic’ microglia (*Est* = 0.042, *t* = 3.43 *p* = 0.00076; Figure 3C). The association remained significant with the addition of covariates for the interval from PET scanning to death and/or PSP pathology stage^28^.

**Figure 3:**
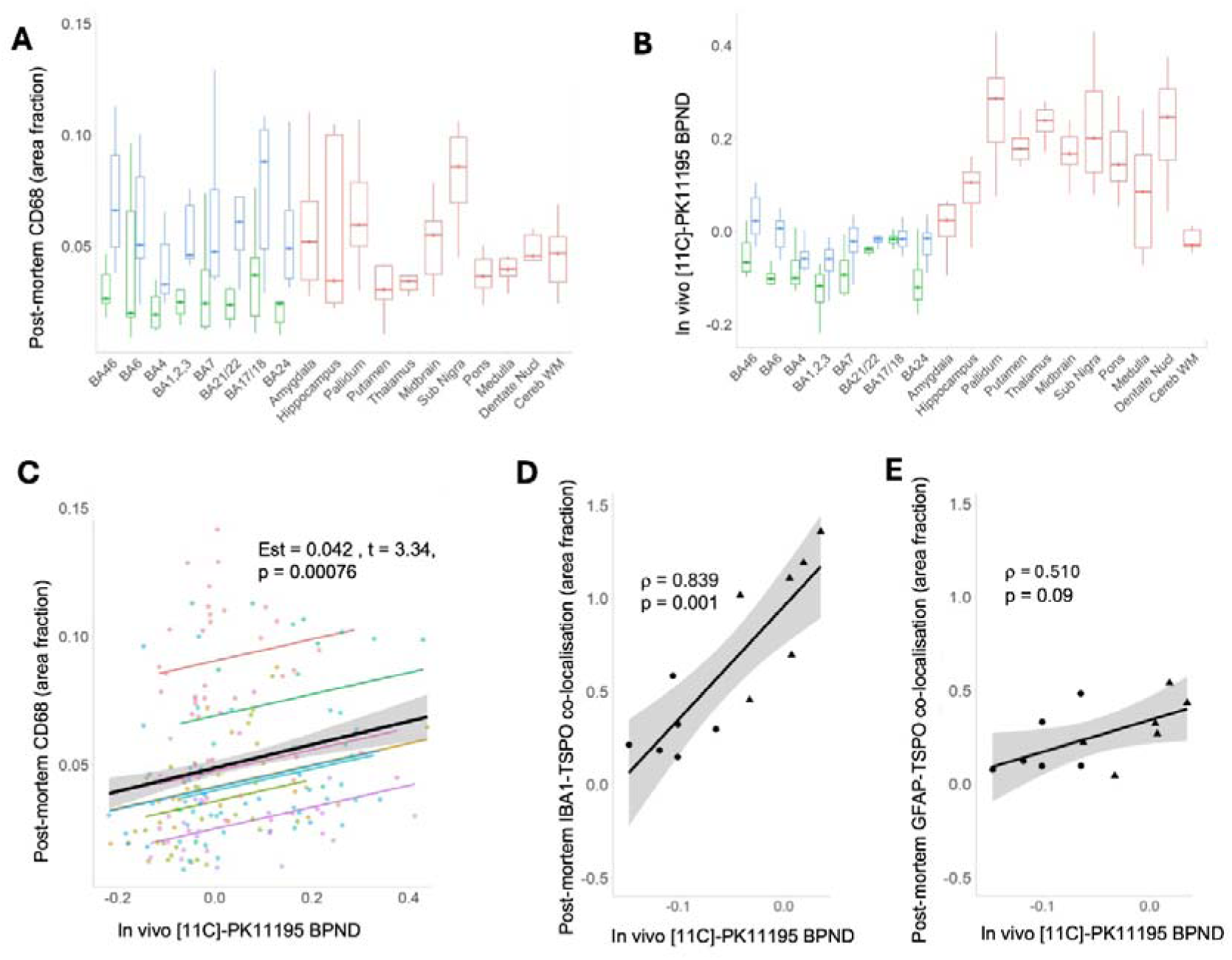
PET to *post-mortem* correlation analysis of [^11^C]-PK11195 BP_ND_ to CD68+ microglia and TSPO-IBA1 and TSPO-GFAP co-localisation. (**A**) Area fraction analysis of CD68+ microglia plotted by region and quantified in white matter (blue), grey matter (green) and subcortical structures (red). **(B)** [^11^C]-PK11195 binding potential (BP_ND_) plotted by region and quantified in white matter (blue), grey matter (green) and subcortical structures (red). **(C)** Association analysis between regional CD68 area fractions and corresponding *ante-mortem* [^11^C]-PK11195 BP_ND_ showing a significant, positive correlation between CD68+ microglia and *in vivo* [^11^C]-PK11195 BP_ND_. **(D-E)** Association analysis between [^11^C]-PK11195 BP_ND_ and TSPO-IBA1 co-localisation area fraction **(D)** and TSPO-GFAP co-localisation area fraction **(E)** in posterior frontal cortex (BA6).

There was a significant positive association between *in vivo* TSPO radioligand binding and microglial TSPO expression in the frontal lobe (*Spearman’s* ρ = 0.839, *p* = 0.001; Figure 3D), whilst an association with astrocytic TSPO expression was not significant (*Spearman’s* ρ = 0.510, *p* = 0.09; Figure 3E).

## Discussion

Our key findings are that (i) despite the multicellular expression profile of TSPO, increased TSPO expression was specifically microglial, and (ii) there was a significant positive association between TSPO radioligand binding during life and *post-mortem* CD68+ microglia, as well as microglial (but not astrocytic) TSPO expression. These findings indicate that TSPO PET in a neurodegenerative primary tauopathy can be interpreted as a microglial-specific neuroinflammatory biomarker. This supports the interpretations of previous PET studies of PSP, reporting increased TSPO radioligand binding^13–15^.

We confirmed that TSPO is expressed in multiple cell types in the human brain, in line with previous reports^26,27^. However, our cell-type specific TSPO expression studies identified microglia as the cell type driving the increase in TSPO in disease. Although there was a trend of increased astrocytic TSPO expression in PSP compared to controls, this did not reach significance. In a cohort of brain donors who underwent [^11^C]-PK11195 PET during life, we compared *post-mortem* quantifications of microglia and cell-type specific TSPO expression with their corresponding *in vivo* TSPO radioligand binding. The TSPO PET binding *in vivo* correlated with phagocytic microglia *post-mortem*, as assessed by CD68 quantification, and did so across multiple cortical and subcortical brain regions. In the frontal cortex, we confirmed the association of *in vivo* TSPO radioligand binding with microglial TSPO expression *post-mortem*, whilst no such association was found with astrocytic TSPO expression. All these findings indicate that although it is most likely that TSPO radioligand binding has potential contributions from the many cell types expressing TSPO, the cell-type mainly driving the increase in TSPO radioligand binding in PSP is microglia. This supports the interpretation of TSPO PET as a microglia-specific neuroinflammatory biomarker in the primary tauopathy PSP.

We recognise that the cellular substrate driving increased TSPO radioligand binding might be disease specific. While the current study reports an increased microglial TSPO expression in PSP, Nutma *et al*^27^ report a seven-fold increase of TSPO+ astrocytes in multiple sclerosis (versus non-lesioned tissue), in addition to more modest increases in microglia and endothelial cells. Garland *et al*^29^ report associations of microglial TSPO with pTau in the later Braak stages of AD, whilst Nutma *et al*^30^ do not observe increased TSPO expression in microglia or astrocytes of AD hippocampi. Together, these reports suggest effects on TSPO expression specific to disease, brain region and disease stage. Further work is required to explore the possibility that the association between microglia and TSPO radioligand binding is PSP/tauopathy-specific, with PET-to-pathology comparisons needed in other neurodegenerative disorders.

Our study has potential limitations. Despite the relatively small sample size, we had adequate power, given (i) the very large expected effect size from previous PET-only reports of PSP, and (ii) the use of a linear mixed effect model for PET-to-*post-mortem* analyses, including all regional data points available for the eight PSP donors. The semi-quantitative nature of immunofluorescence was accounted for in the histological quantifications, using pixel-based colour thresholding to quantify area fraction.

Advantages of using PSP as a demonstrator condition was the clinical and pathological consistency of Richardson’s syndrome, the relatively short interval between PET scanning and brain donation, and unlike Alzheimer’s disease, the absence of a major vascular contribution to pathogenesis. We used the first generation TSPO ligand [^11^C]-PK11195. While second (e.g., [^11^C]-PBR28) and third (e.g., [^18^F]-GE180) generation tracers may offer a more improved signal-to-noise ratio^5^, they are confounded by common genetic polymorphisms that substantially affect binding affinity. They are therefore less suitable for studies of rare diseases, where gene-stratified recruitment would be especially challenging.

In summary, our findings support the use of TSPO radioligand positron emission tomography as a microglia-specific neuroinflammatory biomarker in the primary tauopathy of PSP. Microglial TSPO expression contributes to *in vivo* [^11^C]-PK11195 binding, over and above astrocytic TSPO expression. Given the extensive evidence from PET imaging of increased TSPO radioligand binding in core pathological regions, and its predictive value for clinical decline, we suggest that TSPO PET can be used to quantify microglial-mediated neuroinflammation in primary tauopathies such as PSP, and thereby assist the design and pace of clinical trials with disease-modifying therapies.

## Data availability

Anonymized *post-mortem* and PET data used for this analysis are available on request. Further participant specific information, images or samples can be requested but are likely to require a data/material transfer agreement to adhere to consent restrictions including protection of confidentiality. For the purpose of open access, the authors have applied a Creative Commons Attribution (CC BY) license to any Author Accepted Manuscript version arising from this submission.

## Acknowledgements

We thank our participant volunteers for their participation in this study and brain donation to the Cambridge Brain Bank, and we gratefully acknowledge the participation of all National Institute for Health Research (NIHR) Cambridge BioResource volunteers. We thank the National Institute for Health Research Cambridge Biomedical Research Centre and staff for their contributions, and NIHR and NHS Blood and Transplant. We thank the Cambridge Brain Bank team for their help with tissue provision, the radiographers and technologists at the Wolfson Brain Imaging Centre and Addenbrooke’s Hospital PET/CT Unit for their role in data acquisition, and the East Anglia Dementias and Neurodegenerative Diseases Research Network (DeNDRoN) for help with participant recruitment.

## Funding

This study was supported by Alzheimer’s Research UK East-Network Centre, British Neuropathological Society, Race Against Dementia Alzheimer’s Research UK (ARUK-RADF2021A-010), Addenbrookes Charitable Trust, Medical Research Council (MC_UU_00030/14; MR/T033371/1), Wellcome Trust (220258), Aligning Science Across Parkinson’s disease, the Cambridge University Centre for Parkinson-Plus (RG95450) and the National Institute for Health Research (NIHR) Cambridge Biomedical Research Centre, including their financial support for the Cambridge Brain Bank (NIHR203312). JvdA is supported by a Wellcome Clinical Research Career Development Fellowship (219615/Z/19/Z), a Wellcome Discovery Award (226653/Z/22/Z), and acknowledges core funding from the Medical Research Council (MRC) to the MRC Mitochondrial Biology Unit (MC_UU_00028/8). The views expressed are those of the authors and not necessarily those of the NIHR or the Department of Health and Social Care.

## Competing interests

The authors report no competing interests pertaining to this manuscript. MM has provided consultancy to Astex Pharmaceuticals; JBR provides consultancy with Astex, Asceneuron, Astronautx, Curasen, CumulusNeuro, Prevail, SVHealth; JOB has acted as a consultant for TauRx, Novo Nordisk, Biogen, Roche, Lilly and GE Healthcare and received grant or academic support from Avid/ Lilly, Merck and Alliance Medical. This is unrelated to the current work.

## Notes

### Author Declarations

The research protocols were approved by the National Research Ethics Service's East of England Cambridge Central Committee, and the UK Administration of Radioactive Substances Advisory Committee.

