## Supplementary Material for "*Post-Mortem* validation of *in vivo* 18kDa Translocator Protein (TSPO) PET as a microglial biomarker"

**Detailed Materials and Methods**

**Human brain samples and donors**

Eight PSP subjects (with *ante-mortem* diagnosis of PSP-Richardson’s syndrome) who underwent [^11^C]-PK11195 PET during life^1,2^ donated their brain to the Cambridge Brain Bank. Neuropathological diagnosis of PSP was confirmed in all cases, as per Rainwater criteria^3^, and tau pathology was staged according to Kovacs *et al*^4^. The demographics of the PSP brain donors are summarised in Table 1. Formalin-fixed paraffin-embedded tissue sections from eight cortical and eleven subcortical areas (Brodmann areas BA46, BA6, BA4, BA1,2,3, BA7, BA21/22, BA17/18, BA24, hippocampus, amygdala, putamen, pallidum, thalamus, midbrain, substantia nigra, pons, medulla, dentate nucleus, cerebellar white matter) of eight PSP donors alongside three control cases were provided by the Cambridge Brain Bank (Neuropathology Research in Dementia protocol, Research Ethics Committee reference 16/WA/0240). The control group consisted of age-matched neurologically healthy controls with minimal to mild age-related pathology.

***In vivo* neuroimaging data**

Full details of the imaging protocol and data acquisition are published elsewhere^1,2^. Briefly, patients underwent [^11^C]-PK11195 PET using dynamic imaging for 75 minutes on a GE Advance or GE Discovery 690 PET/CT (GE Healthcare, Waukesha, USA), together with volumetric 3T magnetic resonance imaging (MRI) on a Siemens Magnetom Tim Trio or Verio (Siemens Healthineers, Erlangen, Germany). For each participant, the aligned dynamic PET image series for each scan was rigidly co-registered to the T1-weighted MRI image. Non-displaceable binding potential (BP_ND_) was calculated in cortical and subcortical regions of interest using two atlases: (1) Brodmann areas (www.nitrc.org/projects/mricron) for cortical regions; and (2) a modified version of the n30r83 Hammersmith atlas (www.brain-development.org), which includes brainstem parcellation and the cerebellar dentate nucleus. Supervised cluster analysis was used to determine the reference tissue time-activity curve and BP_ND_ values were calculated in each ROI using a simplified reference tissue model with vascular binding correction.

**Histological staining**

We used DAB-based immunohistochemical staining to visualise expression of TSPO and CD68. Slides were deparaffinized, washed and underwent antigen retrieval at 60°C for 20 minutes with sodium citrate (pH 6.2) for the CD68 and TRIS-EDTA (pH 9.2) for the TSPO protocol. Slides were quenched with 3% hydrogen peroxide and blocked with 5% Normal Horse Serum. Slides were incubated overnight with primary antibodies (TSPO: EPR5384, Abcam, 1:500; CD68: M0876, DAKO, 1:50). Following incubation with secondary antibodies (TSPO: ImmPRESS® HRP Horse Anti-Rabbit IgG Polymer Detection Kit, Peroxidase, Vector Laboratories, MP-7401; CD68: Leica BOND Polymer Refine Detection Kit, DS9800). ABC solution and DAB were used (VECTASTAIN® ABC-AP Kit, Vector Laboratories, AK-5000; DAB Substrate Kit, Vector Laboratories, SK-4100). Slides were counterstained with haematoxylin and then dehydrated and mounted using DPX. CD68 was quantified across all 8 cortical and 11 subcortical regions in grey and white matter separately.

For immunofluorescence co-staining, slides from the BA6 area were baked for two hours and then soaked in xylene, 100%, 96%, 70% and 50% ethanol in series. Antigen retrieval with TRIS-EDTA (pH 9.2) was done at 90°C for 20 minutes, after which blocking with 2.5% BSA for 1 hour was followed by overnight incubation with primary antibodies (IBA1: A82670, antibodies.com, 1:200; GFAP: A83720, antibodies.com, 1:200; TSPO: EPR5384, Abcam, 1:500; CD31: AB9498, Abcam, 1:500). Biotum TrueBlack® Lipofuscin Autofluorescence Quencher (A11055, Invitrogen) was used to reduce autofluorescence and secondaries were added (A11055, Invitrogen: 1:250 for IBA1, 1:500 for GFAP, 1:250 for CD31; A10042, Invitrogen: 1:250 for TSPO). The slides were stained with DAPI and mounted using Fluoromount. Two cases were excluded for GFAP and IBA1 analyses respectively, because the slides did not yield fluorescent signal when stained and scanned.

**Imaging and quantification**

A Leica© SPE Confocal Microscope was used for high magnification images and z-stacks. Whole-slide images were acquired by an Aperio AT2 whole slide scanner (Leica) at x40 magnification for immunohistochemistry slides, and a Zeiss Axioscan Z1 Slidescanner at x40 for immunofluorescence slides.

Area fraction and co-localisation analysis of the IBA1/TSPO and GFAP/TSPO whole slide-scans was performed using a colour-thresholding pipeline in ImageJ. 16-bit images of the white and grey matter ROIs excluding DAPI were used to derive respective RGB colour files, used to quantify colocalisation and total area. Thresholds for each parameter were optimised to the grey and white matter ROIs. Colocalisation colour thresholding was specifically determined as yellow signal i.e., overlapping red (TSPO) and green (GFAP/IBA1) signals.

QuPath was used to quantify CD68+ staining through pixel-classification based analysis in grey and white matter differentiated ROIs. Thresholding for each antibody was then applied to individual images with a gaussian pre-filter, smoothing sigma of 0.5, and a greyscale intensity less than 80. The area fraction was determined as percentage of stained area relative to total tissue area.

**Statistical Analysis**

All cases were blinded until the statistical analysis of data. Non-parametric testing was carried out on all data. Mann-Whitney tests were conducted on the relevant comparisons between control and PSP datasets. The Kruskal-Wallis rank sum test and Dunn’s post-hoc test were applied to the grey matter/white matter comparisons for IBA1-TSPO and GFAP-TSPO area fraction analysis.

A linear mixed effects model was performed to investigate the association between *in vivo* [^11^C]-PK11195 BP_ND_ and CD68+ microglia quantification across all regions. Specifically, CD68 quantification was included as dependent variable and [^11^C]-PK11195 BP_ND_ as fixed factor, while random effects accounted for patient individual variability. As for our previous work^1^, we compared three models using analysis-of-deviance: (1) an initial model with only a random intercept term for patients, (2) a model also including the fixed effect of regional [^11^C]-PK11195 BP_ND_ (x variable) on CD68 quantification (y variable), and (3) a model with both random intercept and random slope terms. Spearman’s correlation analyses were performed to assess the association between the *in vivo* [^11^C]-PK11195 BP_ND_ and TSPO-IBA1 co-localisation and TSPO-GFAP co-localisation area in frontal lobe.
